# Quantitative SARS-CoV-2 tracking of variants Delta, Delta plus, Kappa and Beta in wastewater by allele-specific RT-qPCR

**DOI:** 10.1101/2021.08.03.21261298

**Authors:** Wei Lin Lee, Xiaoqiong Gu, Federica Armas, Franciscus Chandra, Hongjie Chen, Fuqing Wu, Mats Leifels, Amy Xiao, Feng Jun Desmond Chua, Germaine WC Kwok, Shreshtha Jolly, Claire YJ Lim, Janelle Thompson, Eric J Alm

## Abstract

The Delta (B.1.617.2) variant has caused major devastation in India and other countries around the world. First detected in October 2020, it has now spread to more than 100 countries, prompting WHO to declare it as a global variant of concern (VOC). The Delta (B.1.617.2), Delta plus (B.1.617.2.1) and Kappa (B.1.617.1) variants are all sub-lineages of the original B.1.617 variant. Prior to the inception of B.1.617, vaccine rollout, safe-distancing and timely lockdowns greatly reduced COVID-19 hospitalizations and deaths. However, the Delta variant, allegedly more infectious and for which existing vaccines seemed less effective, has catalyzed the resurgence of cases. Therefore, there is an imperative need for increased surveillance of the B.1.617 variants. While the Beta variant is increasingly outpaced by the Delta variant, the spread of the Beta variant remains of concern due to its vaccine resistance. Efforts have been made to utilize wastewater-based surveillance for community-based tracking of SARS-CoV-2 variants, however wastewater with its low SARS-CoV-2 viral titers and mixtures of viral variants, requires assays to be variant-specific yet accurately quantitative for meaningful interpretation. Following on the design principles of our previous assays for the Alpha variant, here we report allele-specific and multiplex-compatible RT-qPCR assays targeting mutations T19R, D80A, K417N, T478K and E484Q, for quantitative detection and discrimination of the Delta, Delta plus, Kappa and Beta variants in wastewater. This method is open-sourced and can be implemented using commercially available RT-qPCR protocols, and would be an important tool for tracking the spread of B.1.617 and the Beta variants in communities.

## INTRODUCTION

The coronavirus disease-2019 (COVID-19) pandemic, caused by the etiological agent SARS-CoV-2, has triggered major public health, economic and psycho-social consequences worldwide (Zoumpourlis et al., 2020). Over the course of the pandemic, new viral genetic variants of SARS-CoV-2 have emerged, yielding variants of concern (VOCs) and variants of interest (VOIs) (Lauring and Hodcroft, 2021). Variants may possess increased transmissibility, more severe disease manifestation, or decrease effectiveness of available medical countermeasures like vaccinations and warrants close monitoring (CDC, 2021). Kappa (B.1.617.1) and Delta (B.1.617.2) variants first emerged in India in October, 2020 (PHE, 2021). The Delta variant has been predicted to be 40-60 % more transmissible than the Alpha variant (Mahase, 2021; PHE, 2021). In comparison to the Alpha variant, the Delta variant is more resistant to vaccine- and infection-induced immunity (Lustig et al., 2021; Bernal et al., 2020), with reduced sensitivity to antibody neutralization (Planas et al., 2021a). The Delta variant has spread rapidly to 104 countries, catalysing waves of COVID-19 infections worldwide (WHO, 2021). In mid March 2021, sequences of the Delta variant with a mutation K417N in the spike protein were detected. This was dubbed the Delta plus (B.1.617.2.1) variant. This K417N mutation is also present in the VOC Beta (B.1.351), which was first discovered in South Africa in 2020 and now reported in 123 countries. While its global prevalence has fallen in the presence of the Delta variant, there remains vigilance on the spread of the Beta variant due to it being more vaccine-resistant than others in circulation (Planas et al., 2021b, Charmet et al., 2021).

The emergence of VOCs and VOIs means that tracking their introduction and spread in populations becomes essential to manage the pandemic. This has mainly been performed via genomic sequencing of clinical samples (Behrmann and Spiegel, 2020). However genomic sequencing of individual clinical samples is expensive and requires specialized infrastructure (Gwinn et al., 2019). Furthermore, for effective surveillance, a significant fraction of infected individuals need to be tested. A companion to clinical surveillance is wastewater-based surveillance, which has been shown to be effective at capturing temporal trends in viral circulation during this COVID-19 pandemic (Hassard et al., 2021; Thompson et al., 2020; Wu et al., 2020a). Current mainstream methods for variant tracking in wastewater rely on enriching and sequencing of the environmental SARS-CoV-2 genome (Napit et al., 2021, Crits-Christoph et al., 2021; Fontenele et al., 2021). This method, constrained by the same bottlenecks of cost and infrastructure requirement as clinical sequencing, is further hampered by challenges in detection of low-frequency variants and is poorly quantitative due to the lack of robust modelling to quantify variant titers (Fuqua et al., 2021; Van Poelvoorde et al., 2021). RT-qPCR-based methods have been developed for variant identification in clinical samples (Clark et al., 2021; Vogels et al., 2021; Wang et al., 2021) but are not yet validated for quantification of variant mixtures, which are expected in wastewater samples. This motivates the development of specialized methods for quantification of variant mixtures in environmental samples such as wastewater (Graber et al., 2021; Yaniv et al., 2021a, 2021b; Wurtzer et al., 2021). While RT-qPCR methods cannot discover new variants or identify variants beyond what they are designed for, they are more sensitive and quantitative, providing a readout from sample to data, in just hours.

In our previous work, we developed and validated Allele-Specific RT-qPCR (AS RT-qPCR) assays for quantitative detection of Alpha variant B.1.1.7 in wastewater, tracking its occurrence over time in 19 communities across the United States (Lee et al., 2021). Here, we develop a similar AS RT-qPCR-based assay for the tracking of the Delta, Delta plus, Kappa and Beta variants by targeting five loci - T19R, D80A, K417N, T478K and E484Q that would identify and differentiate these variants. These assays could be performed as individual reactions or for increased throughput, as triplexes. The AS RT-qPCR assay for variant tracking is easily implementable in the conventional RT-qPCR based surveillance workflow commonly used worldwide.

## RESULTS AND DISCUSSION

Following the methodology established in our previous study demonstrating single nucleotide discrimination for mutations associated with the SARS-CoV-2 Alpha variant (Lee et al., 2021), we developed an Allele-Specific RT-qPCR (AS RT-qPCR) panel for tracking mutations indicative of the SARS-CoV-2 variants Delta (B.1.617.2), Delta plus (B.1.617.2.1), Kappa (B.1.617.1) and Beta (B.1.351) and validated this approach for synthetic mixtures of Beta and Kappa VOC RNA in a wastewater RNA matrix (Delta assay validation is currently in progress, subject to availability of commercial RNA standards). To design allele specific primers we screened a panel of primers targeting mutations characteristic of four variants - Delta, Delta plus, Kappa and Beta, and identified primers targeting five loci - T19R, D80A, K417N, T478K and E484Q as having optimal sensitivity and specificity. These five targets are highly predictive for detection and discrimination of Delta, Delta plus, Kappa and Beta variants among currently circulating strains (**Figure 1**). The Kappa variant can be determined using positive tests for E484Q and the Delta variant by T19R and/or T478K. The Delta plus variant is indicated by additional detection of K417N on top of the ones for the Delta variant. K417N is also present in the Beta variant, which can be indicated by positive tests for D80A.

**Figure 1.**
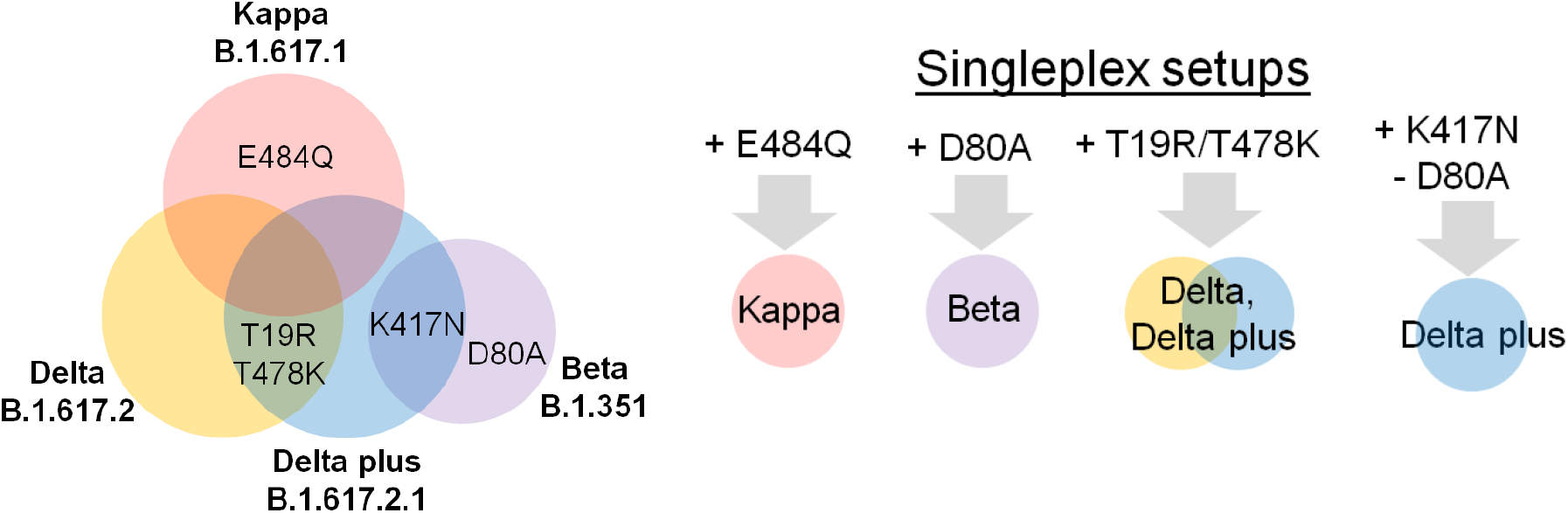
Schematic for deciding which assays to run for identification of Delta, Delta plus, Kappa and Beta variants. **Left**,Venn diagram showing targeted mutations that are representative of each variant. **Right**, Single assays for identification and discrimination of Delta, Delta plus, Kappa and Beta variants. E484Q is unique to the Kappa variant. D80A is unique to the Beta variant. Either T19R or T478K is representative of Delta (inclusive of Delta plus). Delta plus (without Delta) is indicated by subtracting the abundance of K417N (present in both Delta and Beta) by the abundance of D80A (unique to Beta). To exclusively indicate the abundance of the Delta variant, without that of the Delta plus variant, would require subtracting the abundance of the Delta variant by that of the Delta plus. Primers and probes for all assays shown are described in **Table 2**.

We tabulated the presence of these mutations across SARS-CoV-2 variants **(Table 1**) to ensure the absence of cross reactivity with other variants. Both T19R and T478K are unique to the Delta and the Delta plus variant. K417N is present in both Beta and Delta plus. E484Q is unique to the Kappa variant, though variants Beta, Gamma and Eta possess a different mutation, E484K at the same site.

**Table 1.** Characteristic mutations at target loci in Wild Type SARS-CoV-2 and its variants. Mutations to the WT sequence are shown in red (CDC, 2021). (-) denotes no change from wild type SARS-CoV-2.

| WHO name | Scientific name | Amino acid at each loci |  |  |  |  |
| --- | --- | --- | --- | --- | --- | --- |
|  |  | 19 | 80 | 417 | 478 | 484 |
| Wild type |  | T | D | K | T | E |
| Beta | B.1.351 | - | A | N | - | K |
| Kappa | B.1.617.1 | - | - | - | - | Q |
| Delta | B.1.617.2 | R | - | - | K | - |
| Delta plus | B.1.617.2.1 | R | - | N | K | - |
| Alpha | B.1.1.7 | - | - | - | - | - |
| Gamma | P.1 | - | - | T | - | K |
| Epsilon | B.1.427/1.429 | - | - | - | - | - |
| Zeta | P.2 | - | - | - | - | K |
| Eta | B.1.525 | - | - | - | - | K |
| Theta | P.3 | - | - | - | - | K |
| Iota | B.1.526 | - | - | - | - | - |

**Table 2.**
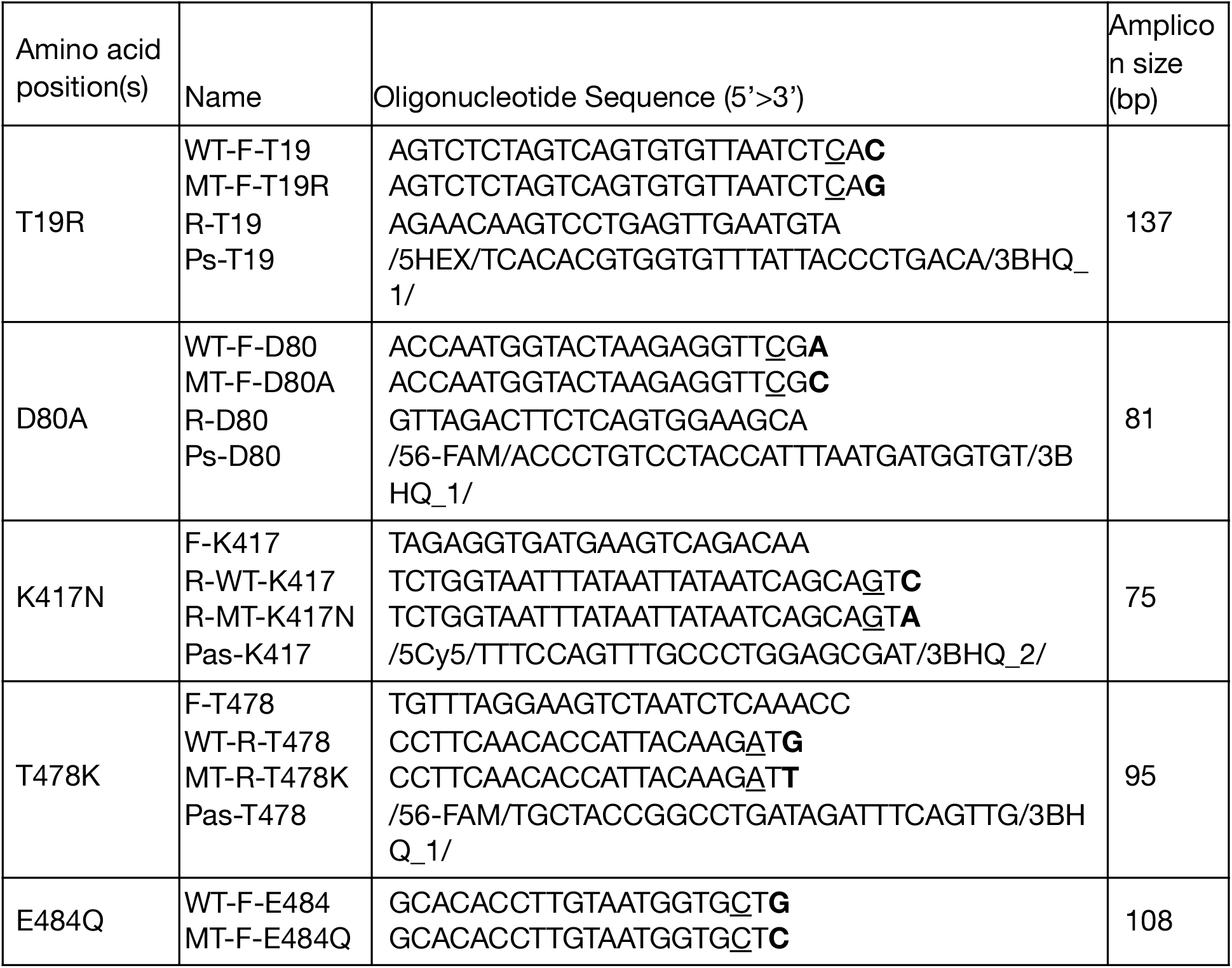
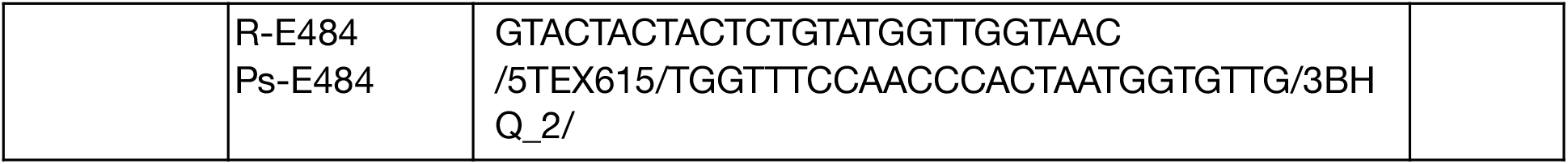
AS RT-qPCR primer sequences developed in this paper for SARS-CoV-2 variants. Allele-specific nucleotides are marked bold. Synthetic mismatches are underlined.

Assays can be run as individual singleplex assays (**Figure 1**) or for increased throughput, as triplex assays depending on the variants to be tracked (**Figure 2**). Triplex setup 1 enables identification of Kappa (E484Q), Delta (including Delta plus) (T19R) and Beta (D80A) variants within the same reaction. This triplex does not enable discrimination between Delta and Delta plus variants. Triplex setup 2 enables identification of Delta (T19R), Delta plus (T19R, K417N) and Beta (D80A, K417N) variants, using D80A to differentiate Delta plus from the Beta variant. K417/N and E484/Q cannot be combined in the same multiplex due to primer interference.

**Figure 2.**
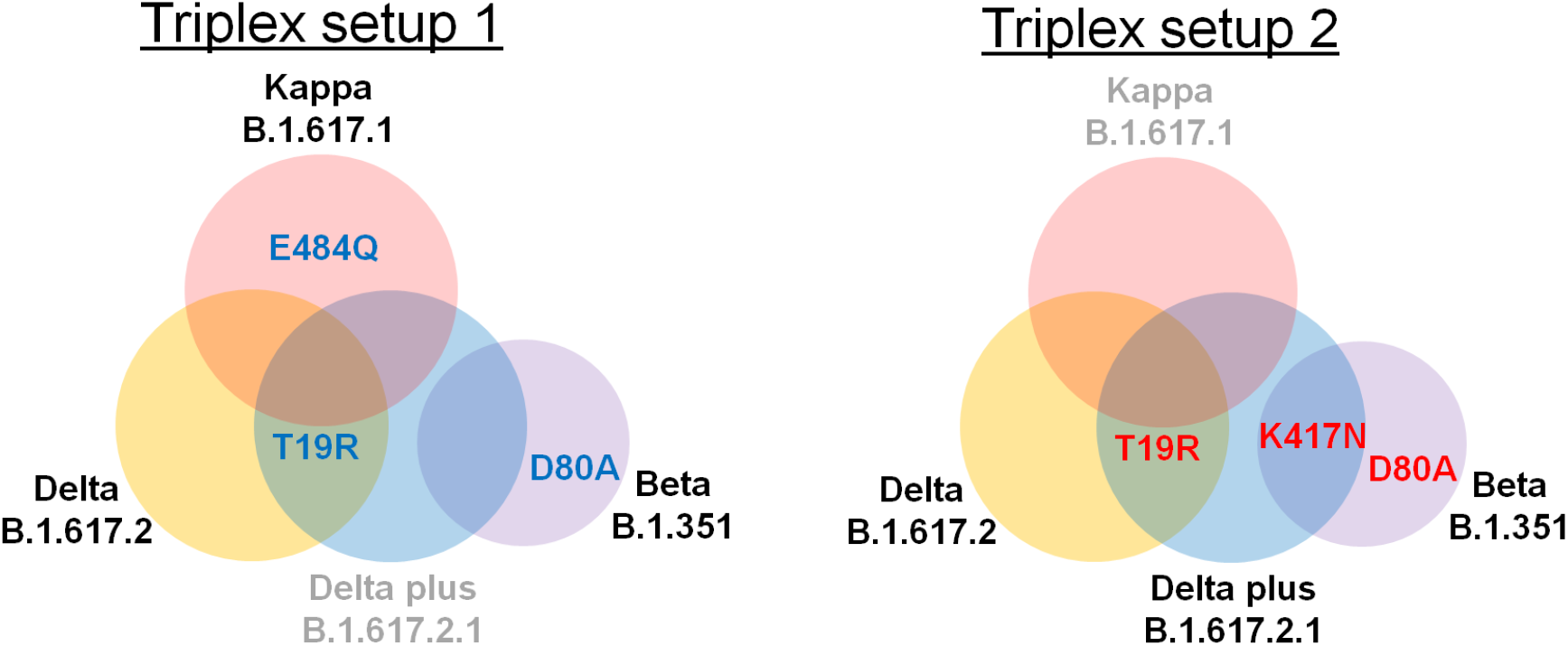
Triplexes for increased throughput of variant detection and quantitation. **Left**, Triplex setup 1 enables identification within the same reaction, the Kappa variant through E484Q, the Delta variant (including Delta plus) through T19R and the Beta variant through D80A. **Right**, Triplex setup 2 enables identification and differentiation of the Delta, Delta plus and Beta variants. The Delta plus variant contains K417N on top of the T19R present in both Delta and Delta plus variants. The Beta variant contains D80A and shares K417N with the Delta plus variant. As such Delta plus is indicated by excluding K417N by D80A. Primers targeting K417/N and E484/Q cannot be combined in the same multiplex due to primer interference.

### Validation of assays that indicate the Beta and Kappa variants

Here we present analytical validation of the sensitivity and specificity of these five pairs of primer-probe sets. Setups are shown in **Table 3**. To improve readability, primers targeting WT are indicated as the amino acid, followed by the position of the amino acid in the protein sequence i.e. T19, D80, K417, T478 and E484 while primers targeting the variants are indicated by a suffix designating the mutant residue i.e. T19R, D80A, K417N, T478K and E484Q. Loci are respectively named T19/R, D80/A, K417/N, T478/K and E484/Q to refer to the loci on both WT and mutant sequences.

**Table 3.** The AS RT-qPCR panel for identification and discimination of Delta, Delta plus, Kappa and Beta variants.

| Locus on the S gene | Target mutation | Variant | Primer-probe set |  |  |
| --- | --- | --- | --- | --- | --- |
|  |  |  | Forward | Reverse | Probe |
| T19/R | T19 | WT | WT-F-T19 | R-T19 | Ps-T19 |
|  | T19R | B.1.617.2 (Delta)<br>B.1.617.2.1 (Delta+) | MT-F-T19R |  |  |
| D80/A | D80 | WT | WT-F-D80 | R-D80 | Ps-D80 |
|  | D80A | B.1.351 (Beta) | MT-F-D80A |  |  |
| K417/N | K417 | WT | F-K417 | R-WT-K417 | Pas-K417 |
|  | K417N | B.1.617.2.1 (Delta+)<br>B.1.351 (Beta) |  | R-MT-K417N |  |
| T478/K | T478 | WT | F-T478 | WT-R-T478 | Ps-T478 |
|  | T478K | B.1.617.2 (Delta)<br>B.1.617.2.1 (Delta+) |  | MT-R-T478K |  |
| E484/Q | E484 | WT | WT-F-E484 | R-E484 | Ps-E484 |
|  | E484Q | B.1.617.1 (Kappa) | MT-F-E484Q |  |  |

We examined the specificity of the AS RT-qPCR assays for Beta and Kappa variant loci D80/A, K417/N and E484/Q by screening them against their respective WT and mutant genome targets in the SARS-CoV-2 RNA (**Figure 3**). Specificity data for T19/R and T478/K is shown in **Figure 4**. Synthetic RNA of the Beta variant was used for D80A and K417N, and Kappa RNA for E484Q. The amplification efficiency of each primer and probe set were between 89.3 to 104% for the correct RNA (i.e. Kappa assay for Kappa RNA, Beta assay for Beta RNA and WT assay for WT RNA). All three assays for mutant sequences and two of three assays for WT sequences were highly specific with cross reactivity only observed at or above 10^3^ copies of non-target sequence per PCR. WT assay K417 had lower specificity indicating cross reactivity at 10^2^ RNA copies of the mutant genotype. All assays were sufficiently specific to support quantification of mixtures at concentrations of SARS-CoV-2 RNA commonly observed in wastewater RNA preparations (Wu et al., 2020a, b, 2021).

**Figure 3.**
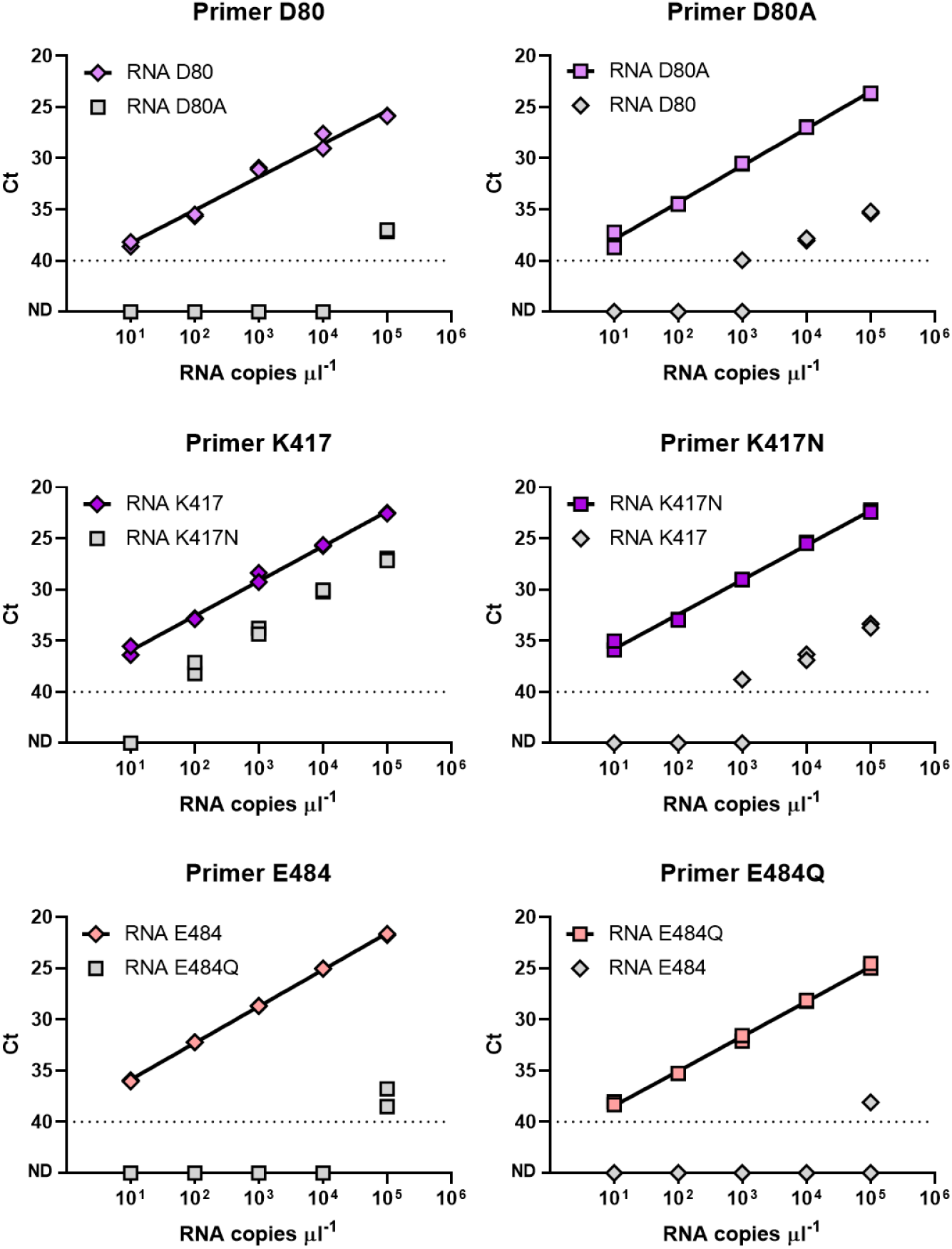
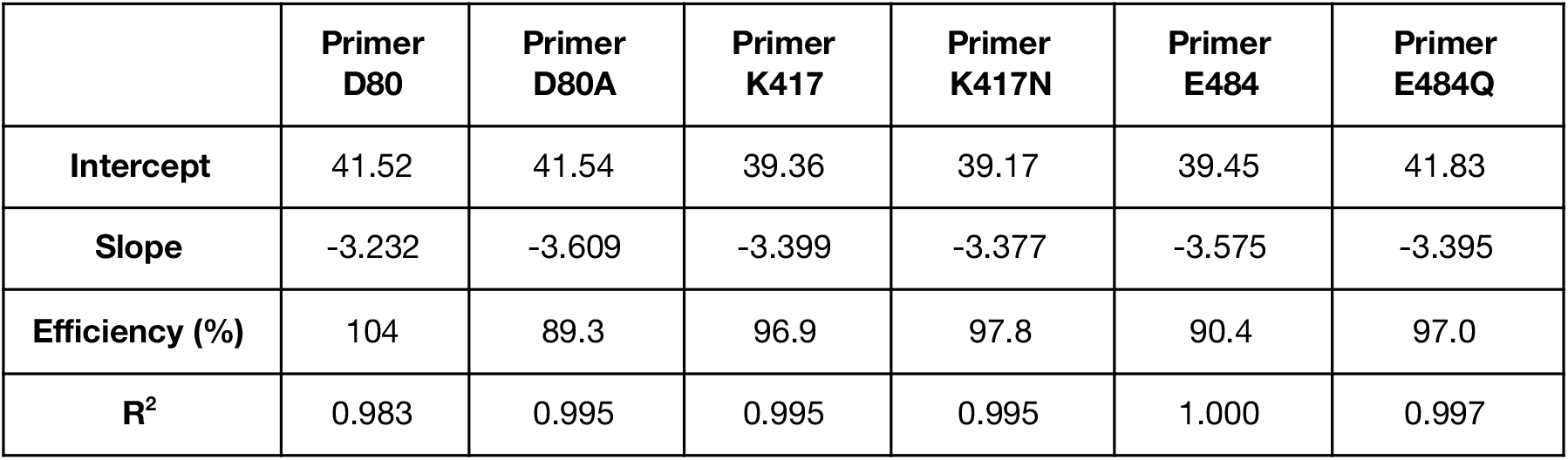
Specificity and cross-reactivity of the three sets of primers against full length synthetic WT, Beta and Kappa variant RNA. Full length synthetic RNA for the Delta and Delta plus variants were not yet commercially available at the time of this work. Primers targeting loci D80/A and K417/N are screened against WT and Beta variant RNA. Primers targeting loci E484/Q are screened against WT and Kappa variant RNA. Exact concentrations of synthetic RNA used were obtained by RT-ddPCR. Colored symbols represent tests against the matching genotype (WT-specific primers to WT RNA and mutant specific primers to mutant RNA) and grey symbols denote tests against RNA of the opposite genotype. Diamonds and squares denote tests using primers designed to target WT and variant RNA respectively. RT-qPCR efficiency and y-intercept cycle threshold (Ct) values were calculated for the primers against their respective RNA target sequences and shown in the table.

**Figure 4.**
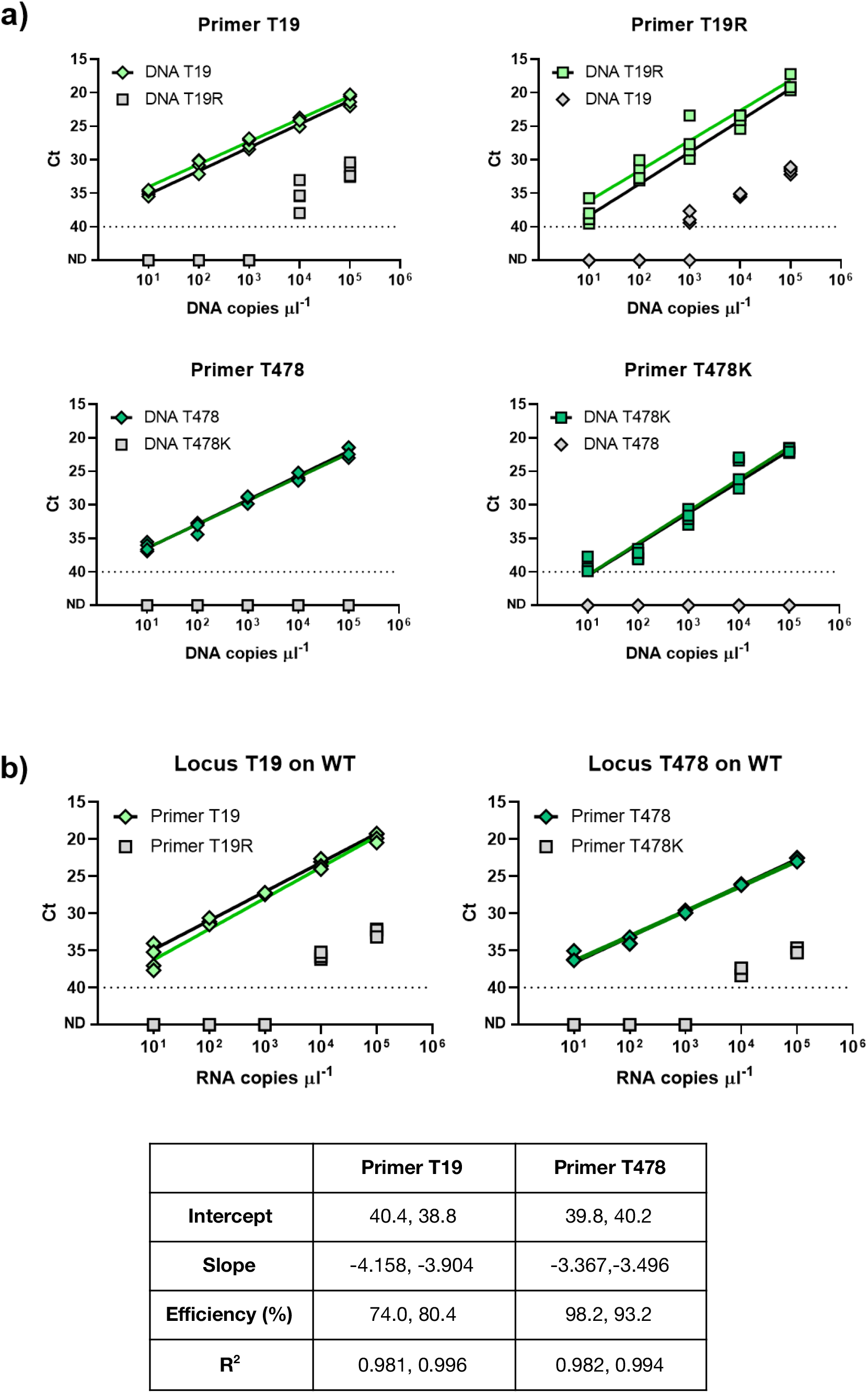
Specificity and cross-reactivity of the two sets of primers against synthetic DNA containing WT or Delta variant sequences. As full length synthetic Delta variant RNA was not yet commercially available at the time of this work, primers targeting loci T19 and T478 are screened against **a**) WT and mutant synthetic DNA and **b**) full length synthetic WT RNA. Exact concentrations of synthetic DNA and RNA were obtained by ddPCR and RT-ddPCR respectively. Data shown reflect two sets of independent measurements taken on different days. Colored symbols represent tests against the matching genotype (WT-specific primers to WT DNA and mutant specific primers to mutant DNA) and grey symbols denote tests against DNA of the opposite genotype. Diamonds and squares denote tests using primers designed to target WT and variant templates respectively. qPCR and RT-qPCR efficiencies and y-intercept cycle threshold (Ct) values were calculated for the primers against their respective RNA target sequences and shown in the table.

### Validation of assays T19R and T478K that indicate the Delta variant

As commercial Delta variant RNA was not available in the laboratory at the time of this work, primers targeting Delta variant loci T19/R and T478/K were validated against synthetic DNA containing WT and variant sequences (**Figure 4a**) (DNA sequences shown in **Table 5**) and full length synthetic WT RNA (**Figure 4b**). The assays targeting T478/K appeared more specific than those targeting T19/R, though all remain discriminant up to at least 10^2^ copies of DNA of the opposite genotype. Screening against WT RNA (**Figure 4b**), the amplification efficiencies for the T19 and T478 primers were between 74.0-98.2%. Mutant primers targeting T19R and T478K did not cross react with up to 10^3^ copies of WT RNA.

### Validation of the singleplex and triplex assays in wastewater RNA

The validation so far suggests most primer-probe sets to be discriminant, with minimal cross-reactivity against at least 10^2^ copies of the opposite genotype. To improve throughput of these assays, we explored which of these primer-probe sets could be multiplexed by combining primers targeting WT loci in the same reaction and primers targeting mutant loci in the other reaction. We found that K417/N and E484/Q cannot be combined in the same multiplex due to primer interference. Further, while primers targeting T478/K were more specific than those for T19/R for identifying the Delta variant, only the latter was incorporated into the triplex assays given overlapping primers, with T478/K being very close in proximity to E484Q.

As such, we developed two triplexes (**Figure 2, Table 4**). The first triplex (Triplex 1) can identify Kappa (E484Q), Delta (including Delta plus) (T19R) and Beta (D80A) variants within the same reaction. Triplex 2 can identify and differentiate Delta (T19R), Delta plus (T19R, K417N) and Beta (D80A, K417N) variants, using D80A to differentiate Delta plus from the Beta variant. Amplification efficiencies between the singleplex and the two triplexes appeared similar for most of the loci (**Figure 5 and 6**).

**Table 4.** Setups for AS RT-qPCR triplexes. All primers and probes are used at a final concentration of 500 nM and 200 nM respectively.

| Triplex 1 for WT | Triplex 1 for Mutant | Triplex 2 for WT | Triplex 2 for Mutant |
| --- | --- | --- | --- |
| <u>Locus T19</u><br>WT-F-T19<br>R-T19<br>Ps-T19 (HEX, BHQ1) | <u>Locus T19R</u><br>MT-F-T19R<br>R-T19<br>Ps-T19 (HEX, BHQ1) | <u>Locus T19</u><br>WT-F-T19<br>R-T19<br>Ps-T19 (HEX, BHQ1) | <u>Locus T19R</u><br>MT-F-T19R<br>R-T19<br>Ps-T19 (HEX, BHQ1) |
| <u>Locus D80</u><br>WT-F-D80<br>R-D80<br>Ps-D80 (Fam, BHQ1) | <u>Locus D80A</u><br>MT-F-D80A<br>R-D80<br>Ps-D80 (Fam, BHQ1) | <u>Locus D80</u><br>WT-F-D80<br>R-D80<br>Ps-D80 (Fam, BHQ1) | <u>Locus D80A</u><br>MT-F-D80A<br>R-D80<br>Ps-D80 (Fam, BHQ1) |
| <u>Locus E484</u><br>WT-F-E484 | <u>Locus E484Q</u><br>WT-F-E484Q | <u>Locus K417</u><br>F-K417 | <u>Locus K417N</u><br>F-K417 |
| R-E484<br>Ps-E484 (TexRed,<br>BHQ2) | R-E484<br>Ps-E484 (TexRed,<br>BHQ2) | R-WT-K417<br>Pas-K417 (Cy5,BHQ2) | R-MT-K417N<br>Pas-K417 (Cy5,BHQ2) |

**Figure 5.**
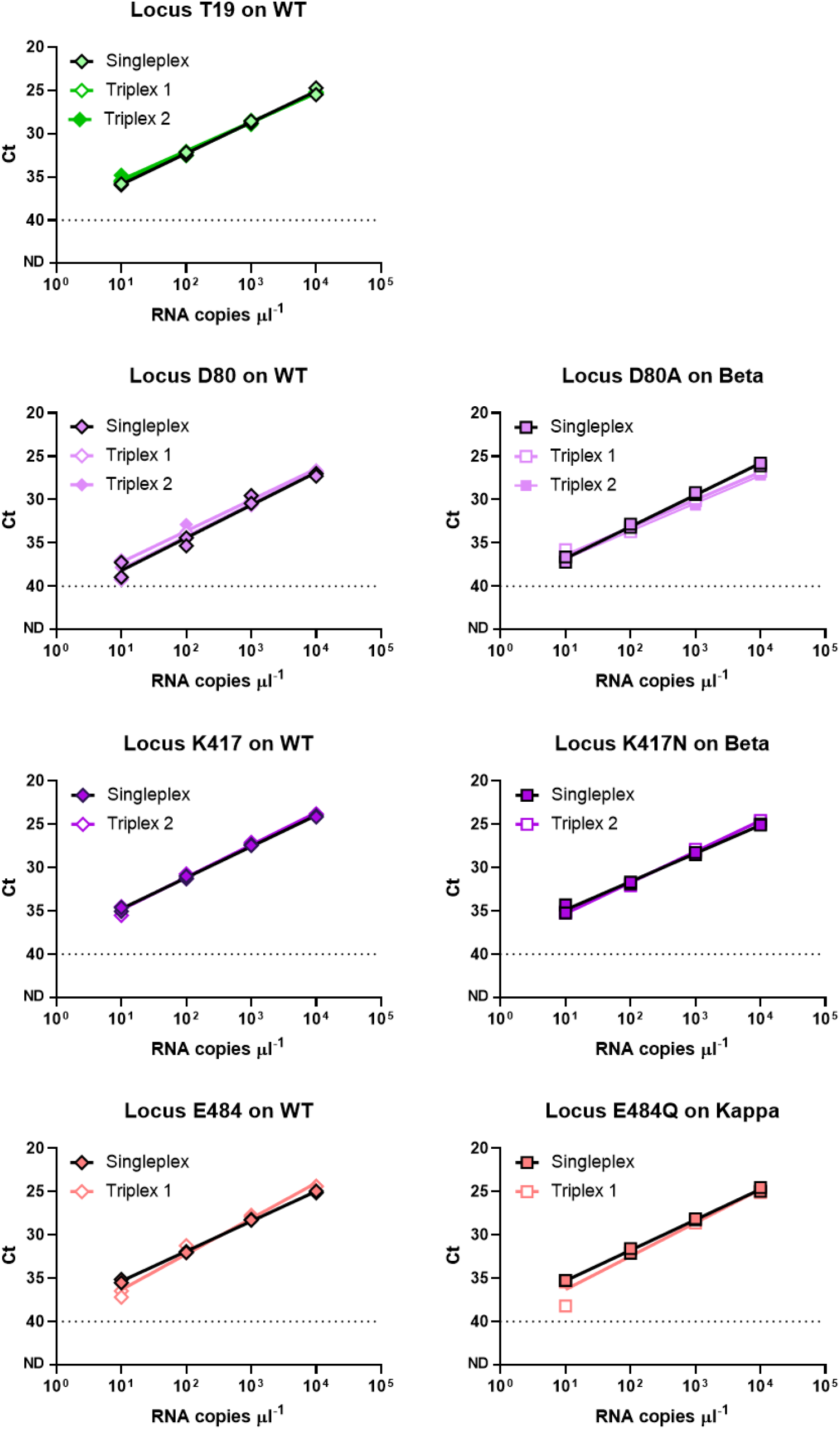
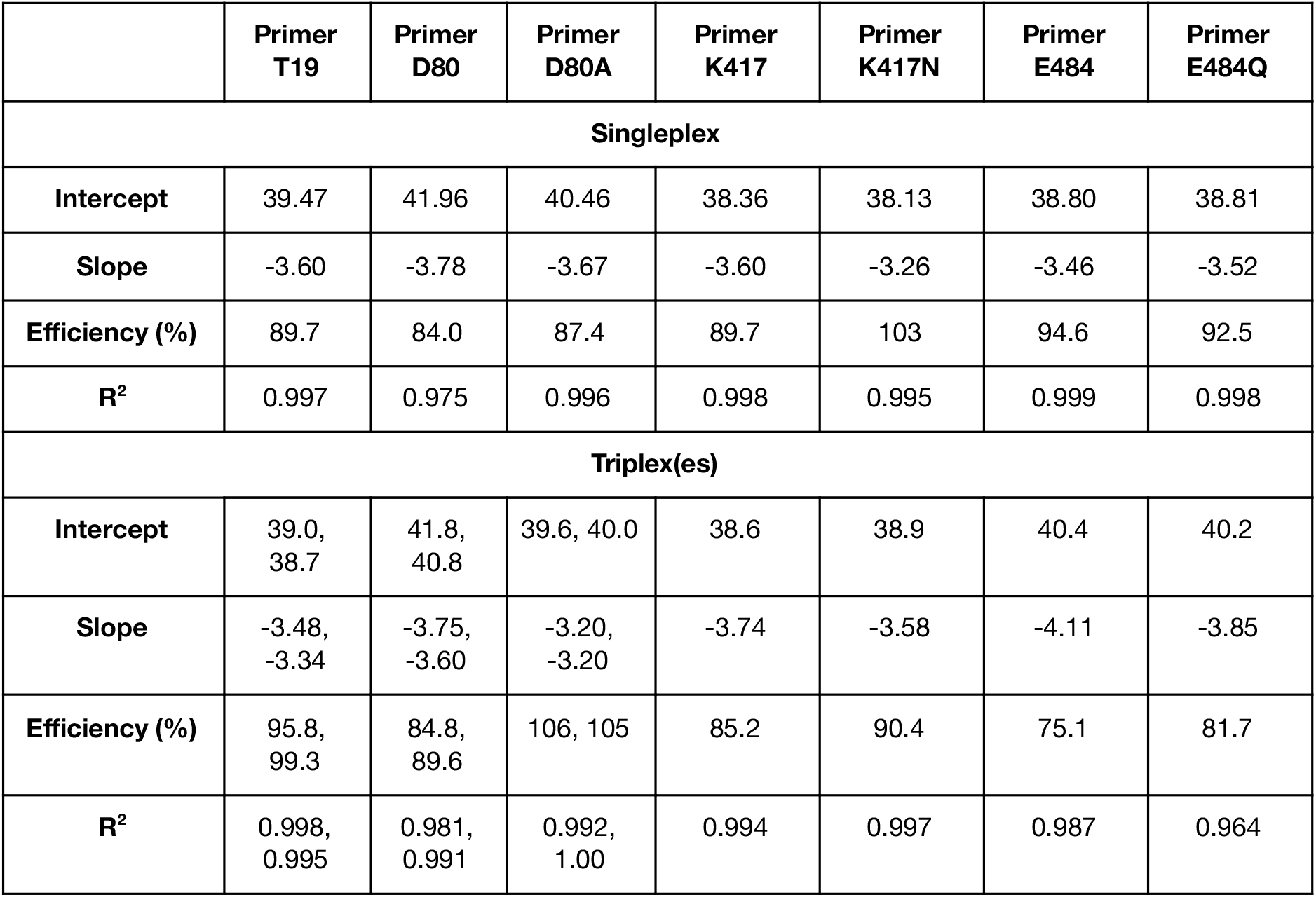
Ct value comparison of singleplex to multiplex AS RT-qPCR using 10-fold serial dilutions of full length synthetic RNA for loci T19, D80/A, K417/N and E484/K. **Left panel**, WT primers are assayed against WT RNA. **Right panel**, primers targeting D80A and K417N were assayed against Beta RNA, and primers targeting E484Q were assayed against Kappa RNA. Diamonds and squares denote tests using primers designed to target WT and variant templates respectively. T19R is not tested against the RNA of the Delta variant and only screened using synthetic DNA templates containing WT and Delta variant sequences, shown in Figure 6. Singleplex and multiplex reactions were performed in the presence of 500 nM of primers and 200 nM of probes. RT-qPCR efficiencies and y-intercept cycle threshold (Ct) values were calculated for the primers against their respective RNA target sequences and shown in the tables. Replicate values represent that of the triplex 1 and 2 respectively.

**Figure 6.**
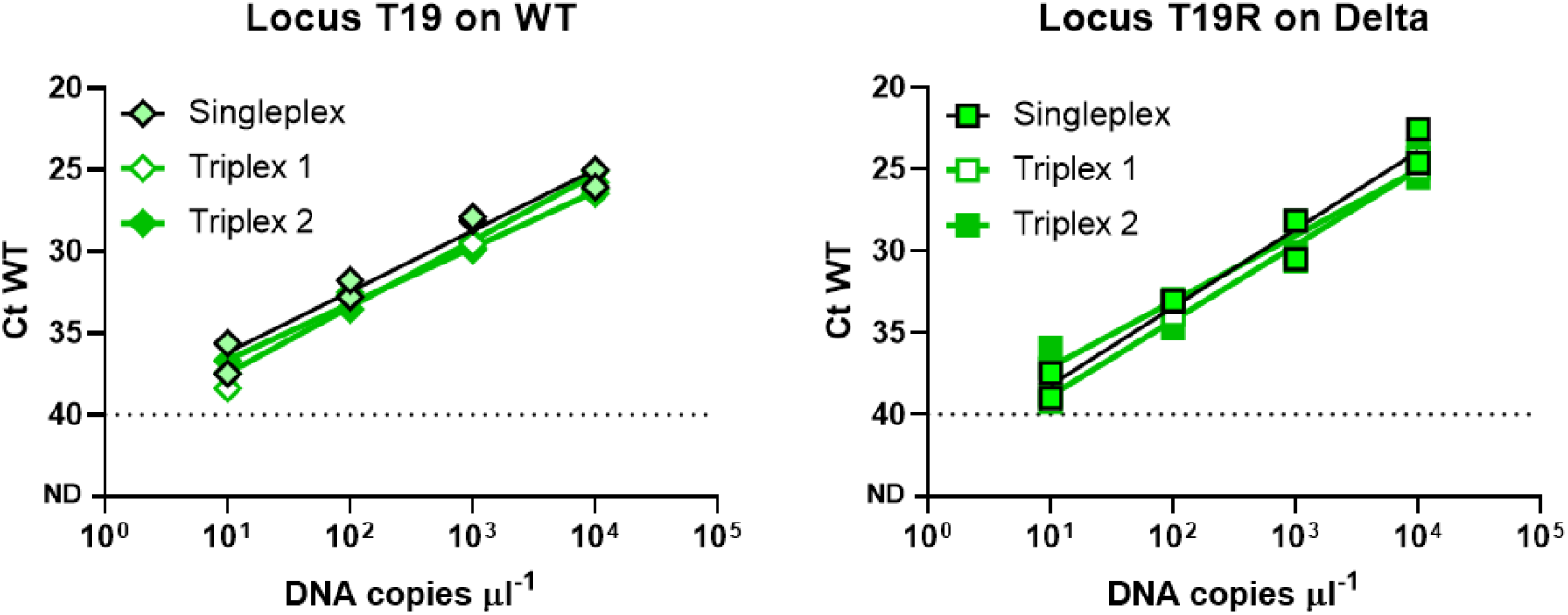
Ct value comparison of multiplex to singleplex AS RT-qPCR using 10-fold serial dilutions of synthetic DNA for loci T19/R. **Left**, WT T19 primers are assayed against synthetic DNA with WT sequences. **Right**, Delta variant targeting T19R primers were assayed against synthetic DNA with Delta variant sequences.

**Figure 7.**
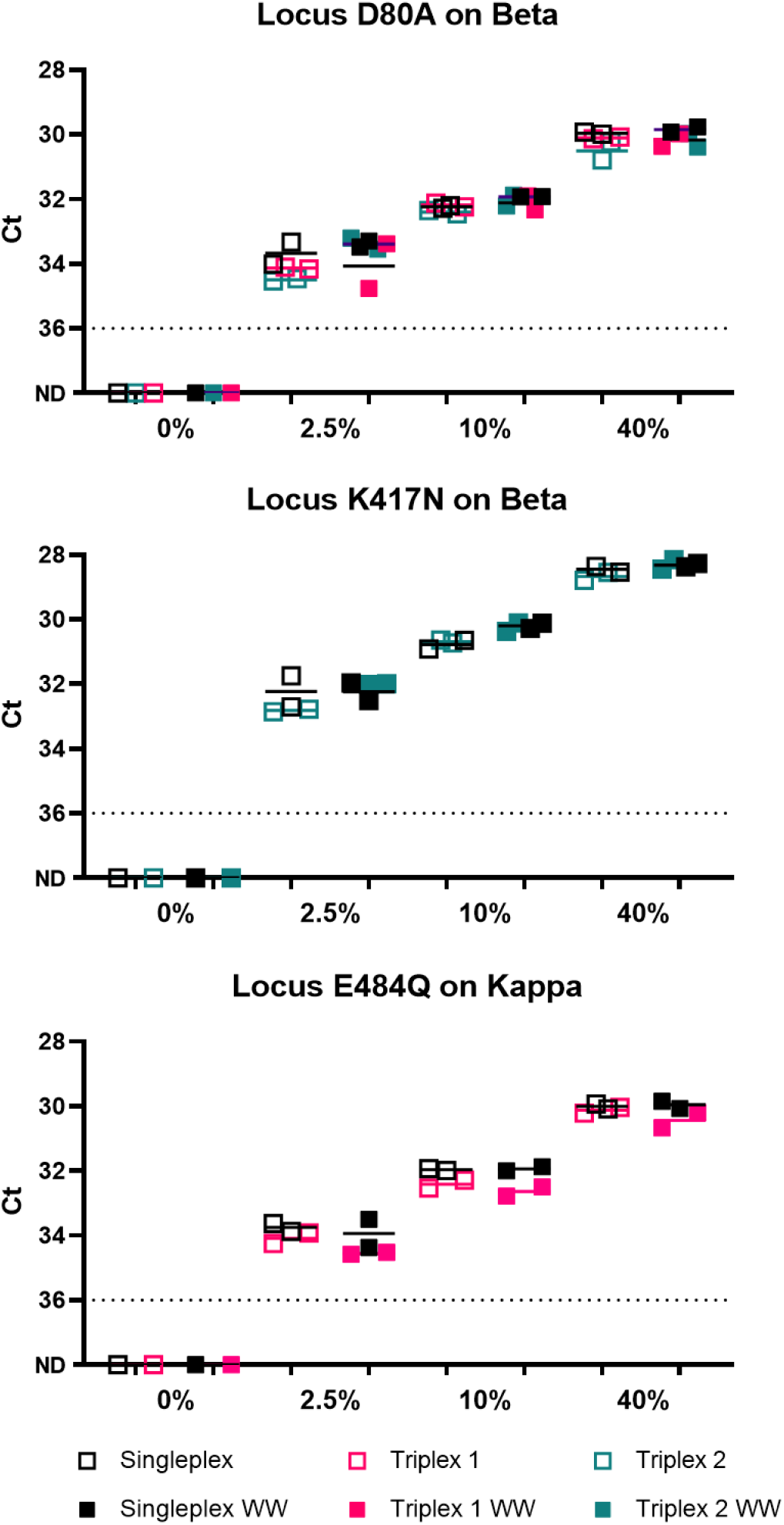
Partial validation of singleplex and triplex assays in a mixture of WT, Beta and Kappa RNA in the presence and absence of wastewater RNA. (Note: Delta variant was not assessed as the synthetic RNA is not yet available in the laboratory). Kappa, Beta and WT RNA were added at 25:25:950, 100:100:800 and 400:400:200 copies per reaction respectively to make up 2.5%, 10% and 40% of Beta and Kappa RNA against a total of 1000 copies of SARS-CoV-2 RNA per reaction. WW refers to assays performed in the presence of wastewater RNA which contained PMMoV at an average of Ct 32–34. Non-specific amplification of delta variant loci was not observed for triplex 1 or 2.

We tested the singleplex and triplex assays in a mixture of WT, Beta and Kappa RNA in the presence and absence of wastewater RNA derived from SARS-CoV-2 negative wastewater. We confirm no amplification with the singleplex and triplex assays targeting Beta, Kappa and Delta variants against wastewater RNA without adding synthetic SARS-CoV-2 RNA. Both the assays (singleplex and triplex) were able to robustly detect synthetic SARS-CoV-2 variant RNA, down to an abundance of 2.5% against a backdrop of 1000 SARS-CoV-2 RNA copies. Similar Ct values were acquired in both assays, when conducted in the presence or absence of wastewater RNA. On average, SARS-CoV-2 RNA derived across wastewater samples tend to be below 10^3^ viral copies per ml, which gives rise to less than 10^3^ viral RNA per RT-qPCR reaction (Wu et al., 2020a, b, 2021). This confirms that the singleplex and multiplex assays are specific and quantitative in wastewater RNA.

## DISCUSSION

The present study presents five assays that could be applied to detecting and differentiating SARS-CoV-2 variants Delta (B.1.617.2), Delta plus (B.1.617.2.1), Kappa (B.1.617.1) and Beta (B.1.351) in wastewater. We followed the screening and validation strategy developed in our previous work on the alpha variant to yield reliable assays that enable discrimination of single nucleotide mutations (Lee et al., 2021). Primer-probe sets were developed for targeting both WT and mutant sequences on loci T19/R, D80/A, K417/N, T478/K and E484/Q. To increase throughput, we propose primer-probe sets that could be combined into triplexes for parallel interrogation of more loci within the same reaction. One of these triplexes enables detection of the Delta, Kappa and Beta variants while the other detects and discriminates Delta and Beta from Delta plus variants. We confirm low cross-reactivity for all the primers to at least 10^2^ copies of RNA of the opposite genotype and similar quantitative performance of singleplex and triplex assays.

Assays for T19/R and T478/K were only validated on synthetic DNA carrying WT and mutant sequences and synthetic WT RNA. Amplification efficiencies were only derived for T19/R and T478/K against synthetic WT RNA and not Delta variant RNA. However WT and mutant assays differ by only a single nucleotide and hybridization kinetics should be similar. Nonetheless detailed analytical validation against the Delta variant RNA will be performed and reported as an update to this preprint.

In this work, we developed primer-probe sets targeting both WT and mutant loci, though variant detection and quantitation would only require performing the reactions targeting the mutant loci. As shown in our previous manuscript (Lee et al., 2021), quantification of both WT and mutant loci in wastewater enables determination of the proportion of WT to variant sequence at each target loci. Validation of the assays reported herein in SARS-CoV-2 positive wastewater is commencing and will be reported when available. This work expands on the utility of AS RT-qPCR for the quantitative detection of variants in wastewater for population-based tracking of SARS-CoV-2 variants.

## MATERIALS AND METHODS

### Assay design

We designed AS RT-qPCR reactions to detect five non-synonymous single nucleotide variants in the spike gene. Primers and probes were designed following our previous work (Lee et al., 2021) and using the Integrated DNA Technologies (IDT)’s PrimerQuest Tool. Target mutations were placed at the 3’ end of either the forward or reverse primer. All primers were designed to have a melting temperature in the range of 59–65°C and the probes in the range of 64–72°C. Probes were designed to anneal to the same strand as the AS primer, with the probe as close to the 3′-end of the AS primers as possible. Guanines are avoided at the 5′-end of the probe. AS primers were designed to include an artificial mismatch near the 3’ terminal nucleotide to improve discrimination. All primers and probes were purchased from IDT (**Table 2**).

### DNA standards and their quantification by ddPCR

DNA standards were synthesised by IDT. Sequences (shown in **Table 5**) were designed to span the primer and probe sets of each assay and then concatenated into a single sequence. This DNA were quantified using ddPCR Supermix for Probes #1863026 (Bio-Rad) following manufacturer’s recommendations. Reaction mixtures consisted of 10 μl of 2× Supermix, 900nM primers, 250 nM probe, 1 μl of DNA, topped up with molecular grade water to a final volume of 20 μl. A Bio-Rad QX200 ddPCR droplet generator (Bio-Rad, USA) was used to convert the reaction mix into droplets. Thermal cycling was performed on a Bio-Rad CFX96 as follows: 10 min at 95°C, followed by 40 cycles of 30 s at 94°C and 1 min at 60°C (ramp rate of 2°C/sec), followed by enzyme inactivation at 98°C for 10 min and holding at 4°C.

**Table 5.**
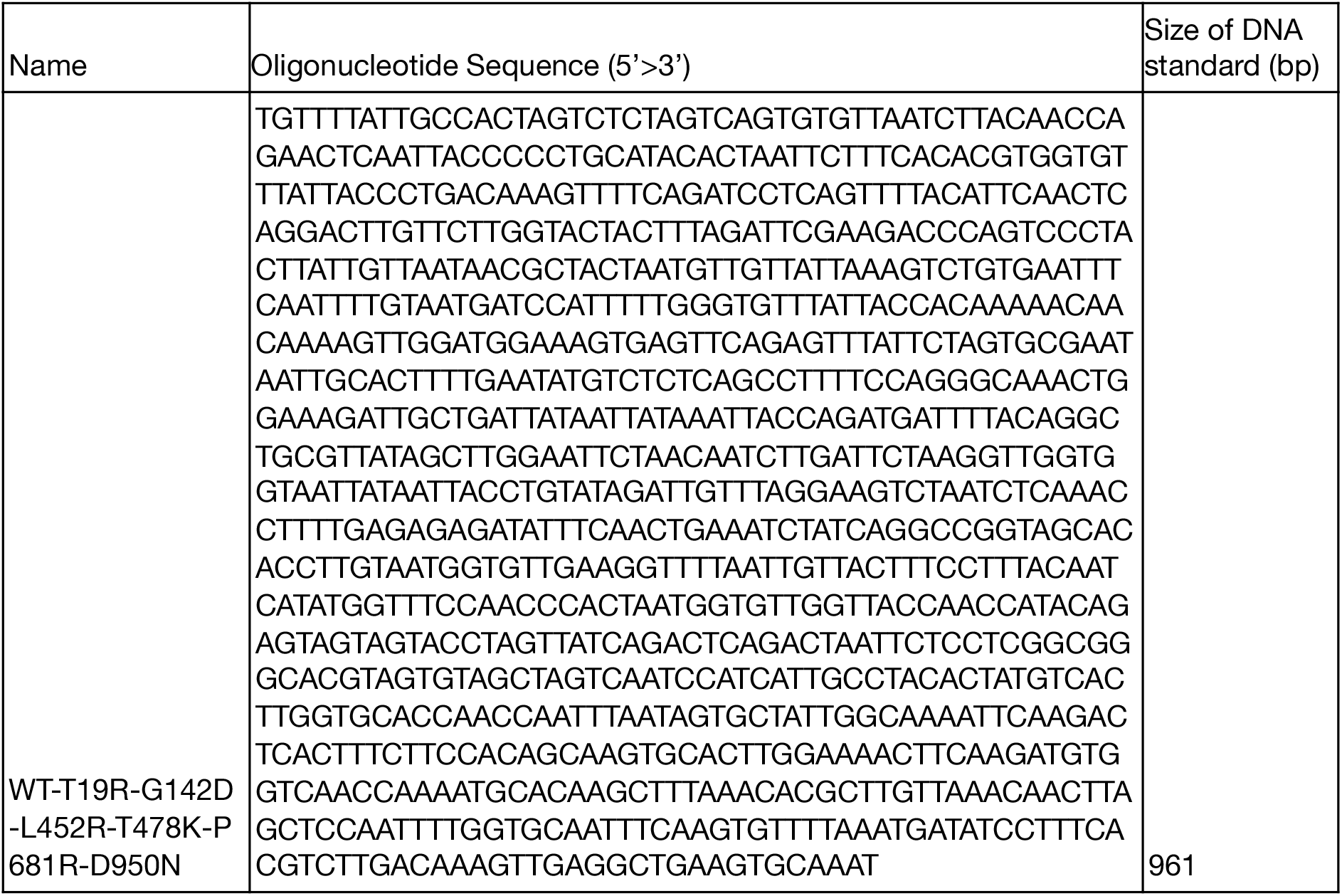
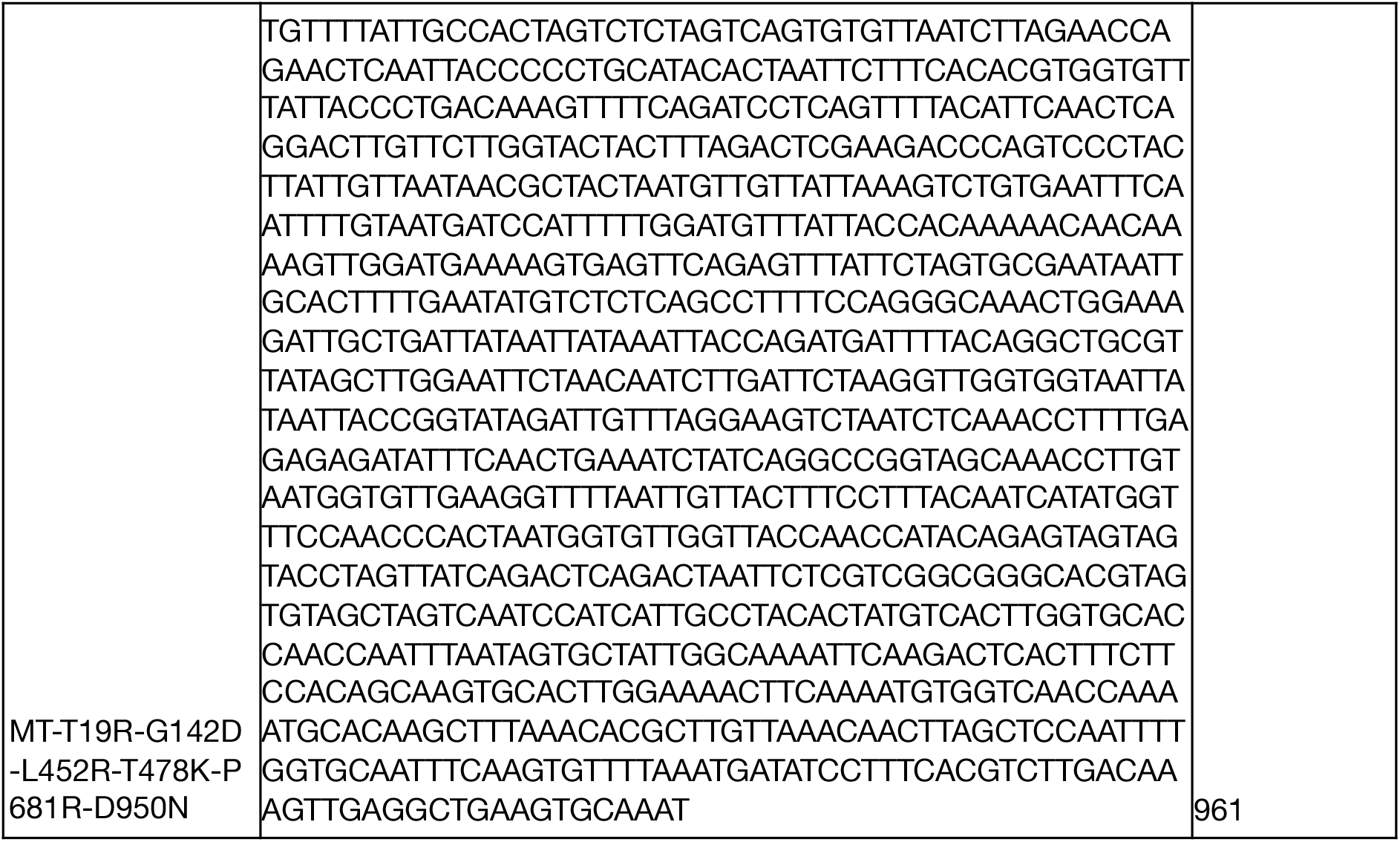
Sequences of DNA standards used for validation of primer sets.

### Analysis of assays against DNA standards by qPCR

qPCR reactions were performed using TaqMan Fast advanced Master Mix (ThermoFisher #4444556) at a final volume of 10 μL in duplicates, according to the manufacturer’s recommendations with a final primer concentration of 0.5 μM and probe of 0.2 μM with 1 μL of DNA template, at 20 s at 95 °C, 40 cycles of 3 s at 95 °C and 30 s at 60 °C) A single reverse or forward primer and probe was used with each set of allele-specific forward or reverse primers (**Table 3**)

### RNA Standards and their quantification by ddPCR

Twist synthetic SARS-CoV-2 RNA Controls, control 2 (Wuhan-Hu-1), control 16 (South Africa/KRISP-EC-K005299/2020), control 18 (India/CT-ILSGS00361/2021), were used as RNA standards, representing WT, Beta (B.1.351) and Kappa (B.1.617.1) variants respectively. RNA standards were prepared as single-use aliquots. Controls 2, 16, and 18 were quantified by digital droplet PCR (ddPCR) to be 4.11 × 10^5^, 8.08 × 10^5^, and 3.24 × 10^5^ copies/μL, respectively. Quantification was performed using One-Step RT-ddPCR Advanced Kit for Probes #1864022 (Bio-Rad) following manufacturer’s recommendations.

### Analysis of assays against RNA standards by RT-qPCR

AS RT-qPCR was performed using the Taqman Virus 1-Step master mix (Thermofisher #4444434) with technical duplicates, at a final volume of 10 µL, according to the manufacturer’s recommendations. For singleplex reactions, a single reverse or forward primer and probe was used with each set of allele-specific forward or reverse primers (**Table 3**). The final concentration of the AS RT-qPCR primers were 500 nM, probe at 200 nM, with 1 µL of template. Multiplex reactions were set up with wild-type targeting primer-probe sets in the same reaction, and mutation targeting primer-probe sets in the other reaction, as shown in **Table 4**, with final concentration of each AS RT-qPCR primer as 500 nM and probe at 200 nM. No template controls were included for each assay and none of them amplified. The reactions are setup using electronic pipettes (Eppendorf) and performed on a Bio-Rad CFX384 real-time PCR instrument under the following conditions, 5 min at 50 °C and 20 s at 95 °C, followed by 45 cycles of 3 s at 95 °C and 30 s at 60 °C.

## Data Availability

Source data will be made available upon request.

## Data analysis

Data was analysed using Microsoft Excel and Graphpad prism. Graphs were presented using Graphpad Prism.

## Data availability

Source data will be made available upon request.

## Contributions

EJA and JT conceptualized the project. WLL designed the experiments. WLL, XG, FA, FC, HC, FW, ML, AX, SJ and CYJL analyzed the data. WLL, FJDC, SJ and CYJL performed experiments. All authors contributed to writing the manuscript. WLL, JT and EJA supervised the project. All authors read and approved the manuscript.

## Acknowledgements

We thank members of the Biobot Analytics, Inc team for helpful discussions.

